# Leveraging multi-source to resolve inconsistency across pharmacogenomic datasets in drug sensitivity prediction

**DOI:** 10.1101/2023.05.25.23290546

**Authors:** Trisha Das, Kritib Bhattarai, Sivaraman Rajaganapathy, Liewei Wang, James R. Cerhan, Nansu Zong

## Abstract

Pharmacogenomics datasets have been generated for various purposes, such as investigating different biomarkers. However, when studying the same cell line with the same drugs, differences in drug responses exist between studies. These variations arise from factors such as inter-tumoral heterogeneity, experimental standardization, and the complexity of cell subtypes. Consequently, drug response prediction suffers from limited generalizability. To address these challenges, we propose a computational model based on Federated Learning (FL) for drug response prediction. By leveraging three pharmacogenomics datasets (CCLE, GDSC2, and gCSI), we evaluate the performance of our model across diverse cell line-based databases. Our results demonstrate superior predictive performance compared to baseline methods and traditional FL approaches through various experimental tests. This study underscores the potential of employing FL to leverage multiple data sources, enabling the development of generalized models that account for inconsistencies among pharmacogenomics datasets. By addressing the limitations of low generalizability, our approach contributes to advancing drug response prediction in precision oncology.

## 1. Introduction

Cancer is a broad group of disorders in which cells develop improperly and may eventually spread to other bodily regions. Due to the varied nature of tumors, treating malignancies remains difficult in modern medicine, making it a serious global public health issue and the second largest cause of death in the United States [1]. Since somatic evolution is the main driving force behind cancer, its genetic underpinnings can be used to define cancer [2]. Clinical decisions can be made based on the types of genetic variants at the individual level since precision oncology aims to tailor medicines to the tumor’s genomic profile. As precision oncology is rapidly developing in clinical practice and is considered the mainstream for treatment, large pharmacogenomics data is considered an important component, which provides information on the drug response for the diverse cancer genomic profiles. The promise of pharmacogenomics data generated in biological settings is to use each patient’s genomic profile to determine the best treatment options for that patient [3]. This makes it possible to create a customized regimen plan when the high cost and lack of free public access to clinical pharmacogenomics databases limit its applicability [4].

The inter- and intra-tumoral heterogeneity is the leading cause resulting in the different responses among cancer patients, which raises notable obstacles in cancer treatment [5]. Thus, drug sensitivity prediction (i.e., drug response prediction) gains popularity in precision oncology by enabling the prediction of the effect of a given drug on a sample (patient or cell line) [6][7][8][9][10][11]. Machine learning (ML) methods have been used to predict drug sensitivity for large panels of cancer cell lines using genomic data and/or drug characteristics. Among these methods, both genomic features [7] [12] and molecular structures/drug descriptors [8][10][11] were also considered. However, because the pharmacogenomics datasets were created for a variety of purposes (such as the investigation of various biomarkers), there are differences in drug responses between various studies on the same cell line using the same drugs. [6][13][14][15][16]. These differences are a result of inter-tumoral heterogeneity [17], experimental standardization [13], and the complexity of cell subtypes. The inconsistency of the pharmacogenomics datasets, which were produced by various studies, places limitations on the drug response prediction based on these datasets. As a result, the predictive model developed based on one dataset has low generalizability and hinders usability when the application is deployed over another dataset. To address the disparities, the uniformity of response measuring techniques is studied to provide better summarization metrics and/or built standardized ones [14][15][16]. For example, Mpindi et al. re-analyzed the dose-response data using a standardized AUC response metric [18]. Hafner et al. proposed GR50, a metric to summarize drug sensitivity which demonstrated better performance in assessing the effects of drugs in dividing cells [19]. Hu, Zhiyue Tom, et al. developed a method called AICM to correct inconsistency between large pharmacogenomics datasets based on cell-wise correlations which also improves drug-wise correlations [20].

Utilizing multiple datasets from various sources can improve prediction performance compared to using single-institution models for common treatment patterns from EHR data [21], demonstrating the potential to be applied to the problem of the disparity between pharmacogenomics datasets in drug response prediction. Here, we adopt Federated Learning (FL), a distributed training of machine learning models without the need to gather data from various sources and compromise privacy, in drug response prediction with high generalizability. On the basis of three pharmacogenomics datasets (i.e., CCLE [22], GDSC2 [23], and gCSI [24]), we examined the proposed model’s ability to handle discrepancy in predicting drug response across cell line-based pair-wise databases. Our model outperforms the baseline methods [21] and the traditional FL method in diverse experiment tests. Extensive experiments are conducted to demonstrate the generalizability of the proposed model. Our novel approach enables a federated model to show the utility of multiple data sources for building generalized models when there are inconsistencies among the multiple datasets. In addition, since the proposed model can be constructed without exchanging data and model, which enables the flexibility for the adoption of our model in different settings (e.g., hospital settings) where the data or model sharing are infeasible.

## 2. Methods and Materials

We have proposed a framework that includes data collection, preprocessing, feature selection, drug representation learning, and model training and testing. Specifically, we built the model based on three well-known cancer cell line publicly available datasets: CCLE [22], GDSC2 [23], and gCSI [24], and trained the model based on overlapping information to learn the disparities in drug responses.

### 2.1. Data

The CCLE [22], GDSC2 [23], and gCSI [24] downloaded from PharmacoDB [25] via the PharmacoGx R package [26] were used as the dataset in the proposed study. Unlike other studies that use only gene expression data [27] or both gene expression data and drug structures [28], we used gene expression data, drug structures, and tissue type information for our prediction models. Our three main data sources are described below.

#### 2.1.1. CCLE

The Broad Institute, the Novartis Institutes for Biomedical Research, and its Genomics Institute collaborated on the Cancer Cell Line Encyclopedia (CCLE) project [22] to characterize a large panel of human cancer cell line models to integrate computational analyses. The CCLE project makes genetic data available to the public and offers analysis and visualization for 1094 cell lines, 25 tissue types, and 24 drugs.

#### 2.1.2. GDSC

The Cancer Genome Project at the Wellcome Trust Sanger Institute (UK) and the Center for Molecular Therapeutics at the Massachusetts General Hospital Cancer Center are working together as part of a Wellcome Trust-funded project called the Genomics of Drug Sensitivity in Cancer (GDSC). The GDSC data contains more than 1100 genetically characterized human cancer cell lines with a variety of anti-cancer therapies. With this vast array of cell lines, scientists are hoping to identify a sizable fraction of the genetic diversity that underlies human cancer and is thought to play a substantial role in the different responses of individuals to various treatments. The GDSC2 data explicitly refers to the recent cell screening technology that GDSC has begun using since 2015 for drug screening.

#### 2.1.3. gCSI

Independent of CCLE and GDSC, the Genentech Cell Line Screening Initiative (gCSI) was started to address concerns about inconsistencies among extensive pharmacogenomic research. In gCSI, Genentech independently assessed how 788 cancer cell lines responded to a subset of the drugs (44) on 27 tissue types examined by GDSC and CCLE.

### 2.2. Data Processing

#### 2.2.1. Overlapping data

There were 706, 599, and 566 overlapping cell lines for CCLE-GDSC, CCLE-gCSI, and gCSI-GDSC respectively. Similarly, there were 12, 8, and 26 overlapping drugs for CCLE-GDSC, CCLE-gCSI, and gCSI-GDSC respectively. Therefore, many drug-cell line pairs were tested in pairs of studies. The overlapping number of samples (drug-cell line pairs) was 3482 for CCLE-GDSC, 4239 for GDSC-gCSI, and 1333 for CCLE-gCSI.

#### 2.2.2. Combining datasets

For this study, we needed to merge gene expression data, drug structure data, tissue type data, and response data. 3.2.1, 3.2.2, and 3.2.3 show how we got embeddings for gene expression data, drug structure data, and tissue type. For each response sample, we concatenated the embeddings followed by the response score to create the final datasets for each of the three source datasets (CCLE, GDSC, and gCSI). We discarded rows with any missing values in the datasets.

#### 2.2.3. Gene Selection

We filtered the genes based on the gene sets used in the Broad L1000 project [29], which were found sufficient to predict the transcriptome change upon drug treatment. Additionally, we further reduced the number of genes based on mutual information. In practice, we calculated mutual information with the sklearn package [30]. We performed a different set of experiments with non-overlapping cell-line and drug pairs between different pairs of datasets (e.g. CCLE-GDSC) where we used all 902 common genes between the L1000 genes and the datasets.

#### 2.2.4. Drug Embedding

To use the knowledge from drug structures for drug response prediction, we collected Simplified Molecular Input Line Entry System (SMILES) codes for all drugs in all three datasets from PubChem using the python package PubChemPy [31]. SMILES is used to convert the three-dimensional structure of a chemical into a string of symbols that computer programs can easily comprehend. However, we needed vector representations of drugs instead of strings for the predictive model. Therefore, we used Mol2Vec [32] to find embeddings of 300 dimensions for each drug.

#### 2.2.5. Tissue type

The tissue type information was included as suggested by the study[9], which shows that non-solid tissue types influence the performance of models. We found that there are 21 cell lines common in all three datasets after all the preprocessing. We used one-hot encoding for tissue types for the model.

### 2.3. Models for drug sensitivity prediction

We used the gene expression, embeddings of SMILES codes, and tissue type of the cell line to predict the sensitivity score. Our model can be represented as:

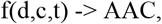

where *d* is an n-dimensional vector representing drugs (i.e., SMILES), *c* is an m-dimensional vector for gene expression of a cancer cell line, *t* is a tissue type, and AAC is the drug sensitivity score.

We chose AAC instead of the well-known IC50 metric as our drug sensitivity measure because models trained to predict AAC outperform those trained to predict IC50 [9].

We propose a novel method for predicting the drug sensitivity of a drug-cell line pair in one source by gathering indirect knowledge from a separate source. Let’s think of Sources 1 and 2 as two distinct pharmacogenomic studies with limitations on data sharing. We expect that some samples from the two sources will always overlap. That implies that some drug-cell line combinations are examined in both studies. We have two local models, m1 and m2, for the two sources. From the embeddings we produced in Section 2.2.3-2.2.4, we presume that m1 and m2 are models which predict drug sensitivity. Both m1 and m2 can be considered black box machine learning regression models. However, we consider that both are similar models. This implies that both m1 and m2 will be neural networks if m1 is a neural network. The predicted values of models m1 and m2 are used to fit models m3 and m4. In the case of m3, the predicted values of m1 serve as the independent variable and those of m2 serve as the response variable. In the case of m3, the predicted values of m1 serve as the response variable and those of m2 serve as the independent variable (Figure 1).

**Figure 1.**
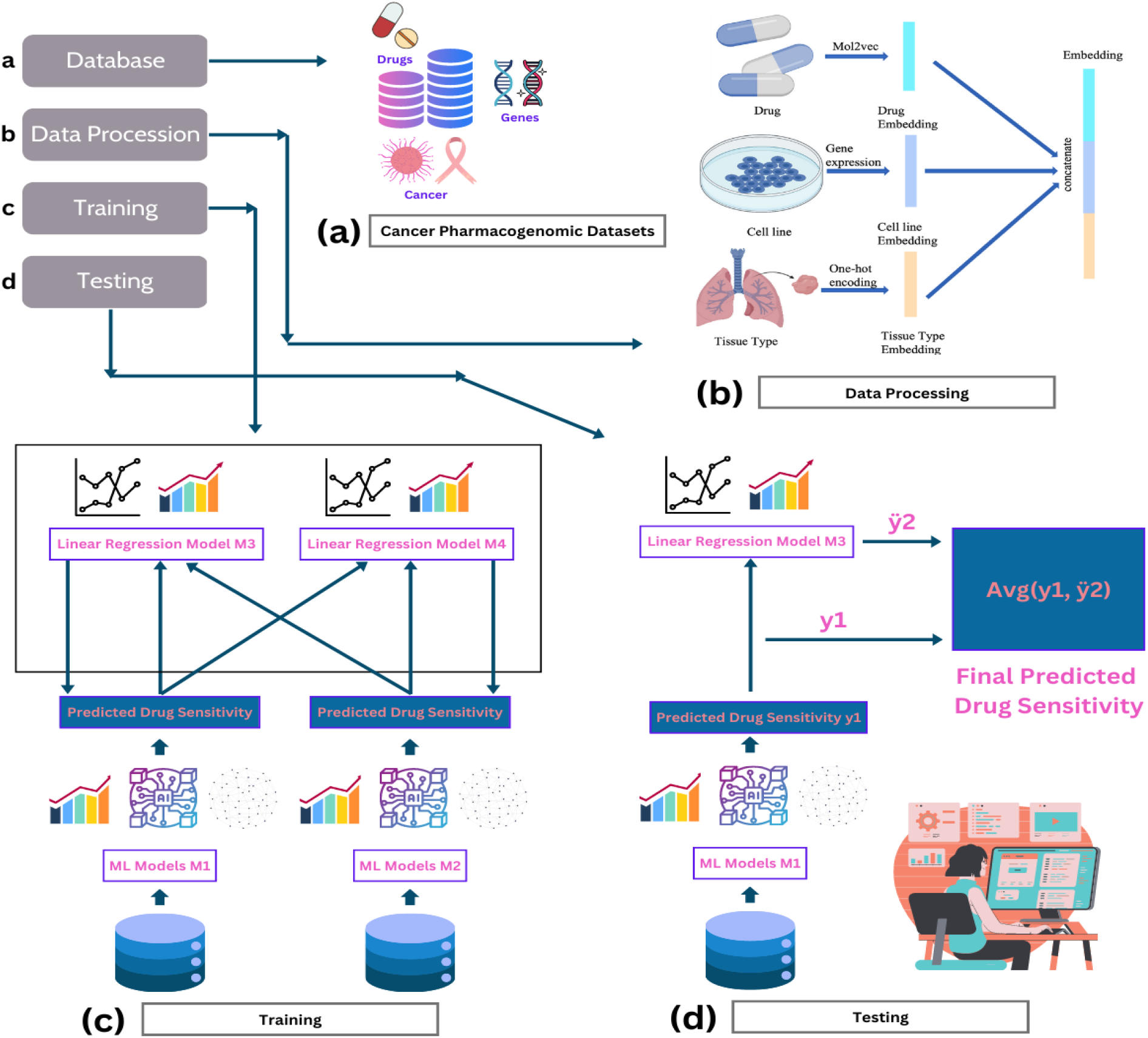
The proposed framework in drug sensitivity prediction. (a) The drug, cell line, and sensitivity score were obtained from the three main datasets: GDSC, CCLE, and gCSI. (b) The process of generating embeddings from drug structures and cell lines, (c) Training process, (d) Testing/inference process only shown for source 1 (model 1), and model 2 follows the same method.

**Figure 2:**
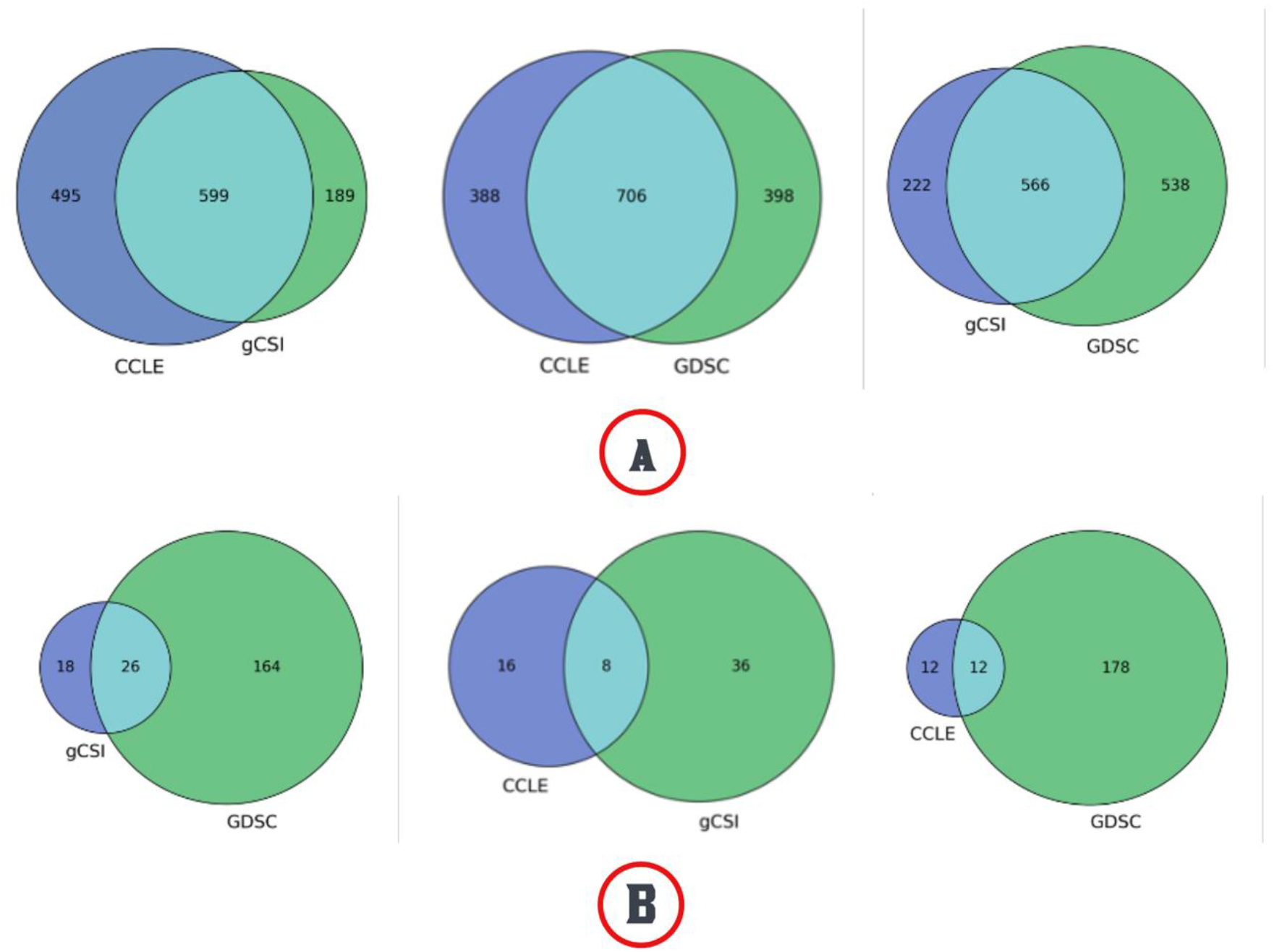
Overlapping data among CCLE, GDSC, and gCSI. (A) Pairwise cell line data overlap, and (B) Pairwise compound data overlap.

**Figure 3:**
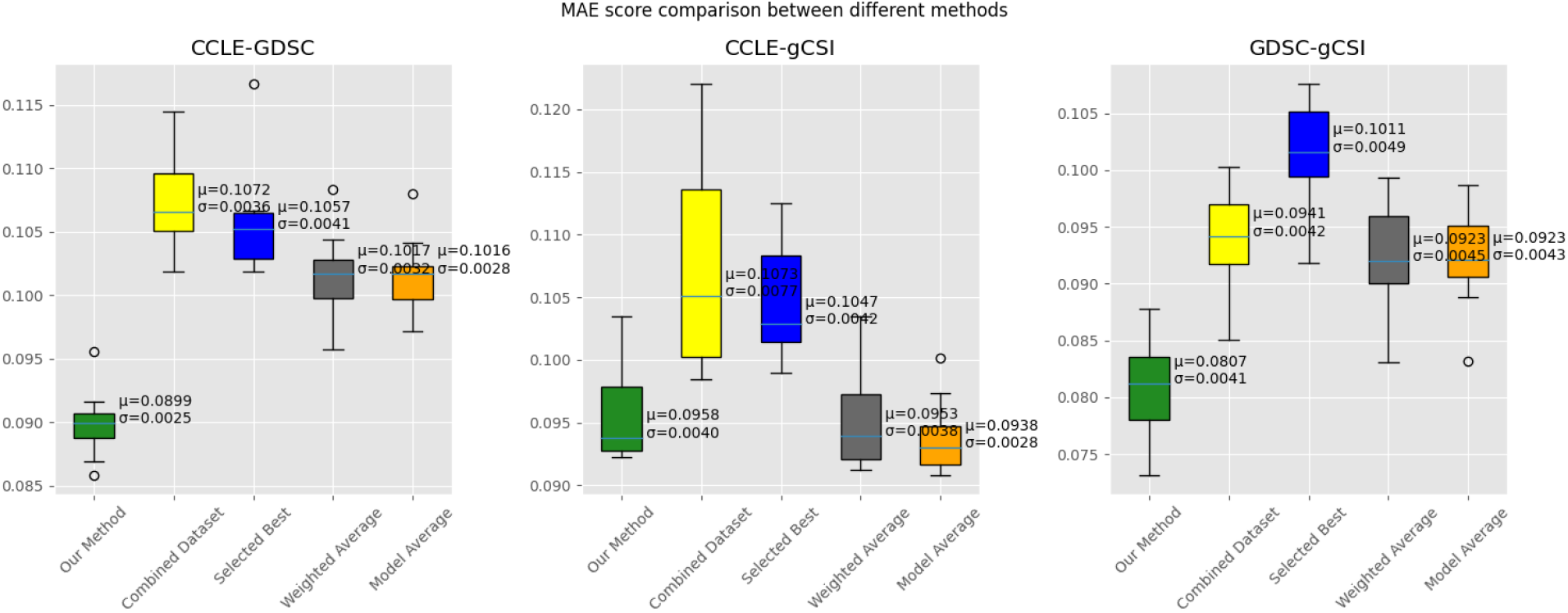
Cross-validation results on common drug-cell line pairs

We used federated learning to our advantage, which enabled us to predict the drug sensitivity of one data source based on training on both data sources. In the event that one data source is not big enough to train an algorithm, this strategy may be highly useful. We could also easily train the small dataset by combining it with another large dataset. We tried this method on various regression techniques (ridge and linear regression) and machine learning algorithms (neural network) to check its robustness.

### 2.4. Experiment design and evaluation

#### 2.4.1. Baseline models in comparison

Inspired by [21], which leverages three evaluation approaches for multi-source EHR data for predicting pharmacotherapeutic outcomes of type 2 diabetes, we compared our model with the three baseline approaches: Selecting Better, Result Average, and Combining Data. We added another approach of Model Average with our proposed model to handle the inconsistency between data from more than one source.

Selecting Better (SB): We trained a model m1 on the training set of the first dataset and a model m2 on the training set from the second dataset. During inference, we tested both models on the test data and reported the model that has higher scores between m1 and m2. We do this for all three datasets (CCLE, GDSC, and gCSI).

##### Result Average (RA)

This is a simpler version of the weighted average method [21]. We only have single correspondence between two datasets for a single drug-cell line pair if it is in the overlapping region of the two datasets. Therefore, we used simple pointwise averaging of the test results from models m1 and m2. That means, for example, while dealing with CCLE and GDSC, we trained m1 on the training data of CCLE and trained m2 on the training data from GDSC. While testing, we collected the predicted AAC for each corresponding pair of samples and averaged those to get the final prediction if the test sample is from the overlapping part of the datasets. Otherwise, only a single prediction value was delivered as output for a non-overlapping sample based on which study it belongs to.

##### Combining Data (CD)

Here we combined the training data from both datasets and train a model m on the combined data. While testing, we used model m to predict the AAC scores of the test data.

##### Model Average (MA)

Like RA and SB, we trained two models m1 and m2 on the corresponding training data from two different datasets. We then averaged the model weights and biases to get a final model. This final model is then used to predict the AAC scores for the test data.

#### 2.4.2. Evaluation metrics

As the task is a regression task where the labels are real values, we computed Mean Square Error (MSE), Mean Absolute Error (MAE), and R-squared (R2) to evaluate model performances. For simplicity, we reported MAE in the main manuscript and put the result of MSE and R2 into the supplementary files.

### 2.5. Evaluation tasks

To evaluate our method, we perform the following tasks.

#### Task 1-Performance on overlapping samples

We did 10-fold cross-validation on the overlapping parts of the data pairs. Each sample in Source 1 has a corresponding sample in Source 2 (Figure 1(b)). We divided data in Source 1 and Source 2 into 10-fold in such a way that all the corresponding samples are in the same fold for both datasets. That means all training samples in Source 1 had a corresponding sample in the training sample of Source 2 which is treating the same cell line with the same drug. A similar setting is maintained in the test set naturally.

#### Task 2-Effect of an increasing number of non-overlapping samples

For this part, we only used the CCLE-GDSC pair. We did 10-fold cross-validation as before. There are 3482 overlapping samples. We kept a certain percentage of non-overlapping samples of one dataset constant and changed the size of the other dataset. For example, we kept 100% of CCLE and then used three different sizes of GDSC: 100%, 70%, and 50%, and vice versa. We then combined overlapping and non-overlapping samples for the three sub-experiments. The models m1 and m2 were trained on training datasets of the corresponding datasets at each time of the 10-fold cross-validation. However, models m3 and m4 were trained on the overlapping parts of the training datasets. Figure 5 shows MAE scores for all 4 baselines with an increasing amount of non-overlap data.

**Figure 4:**
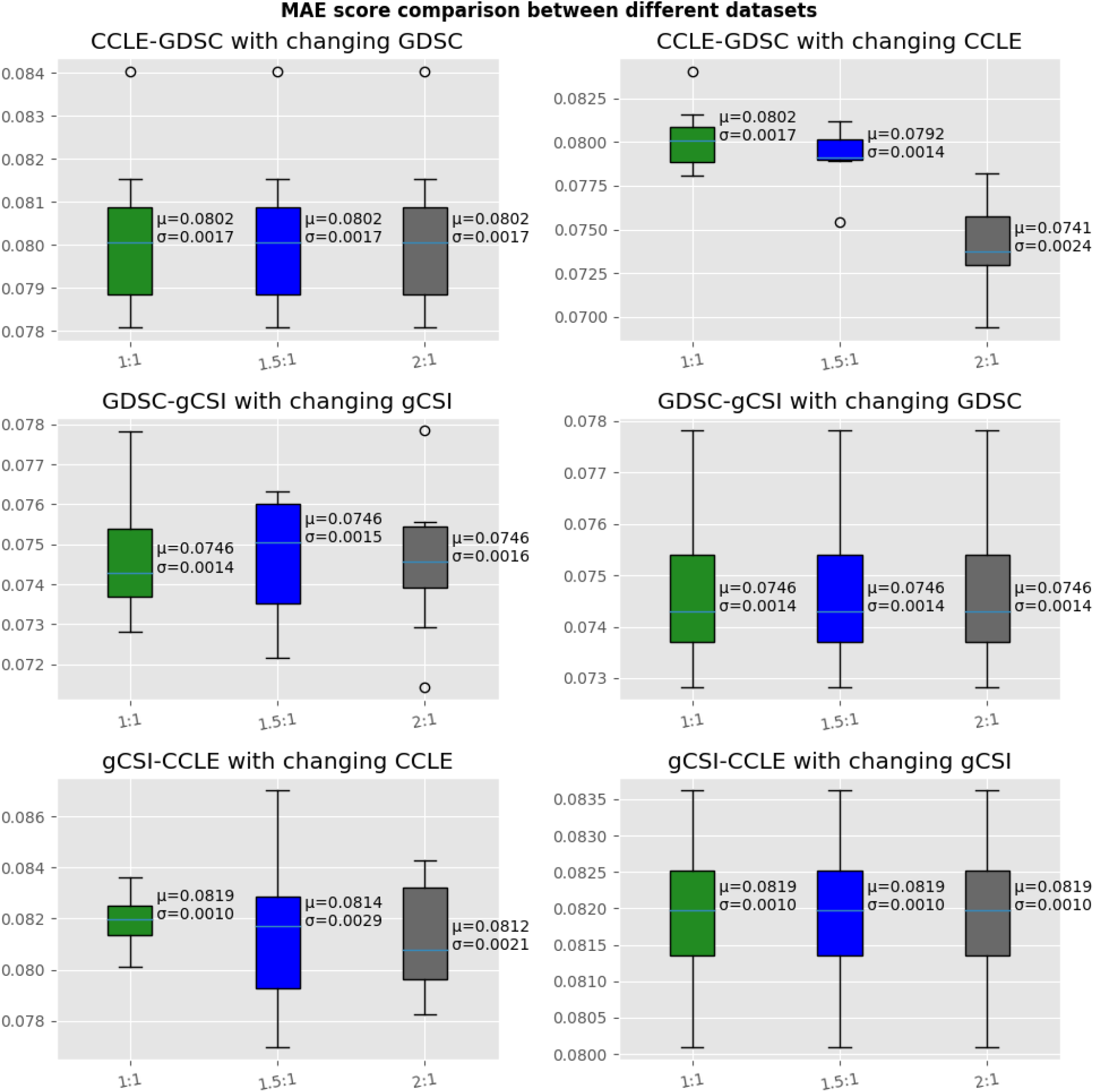
Cross-validation results on CCLE-GDSC with different numbers of non-overlapping samples (left to right). Please note that for gCSI-CCLE with changing gCSI, we could not calculate 1.5:1 and 2:1 results due to inconsistent data.

**Figure 5:**
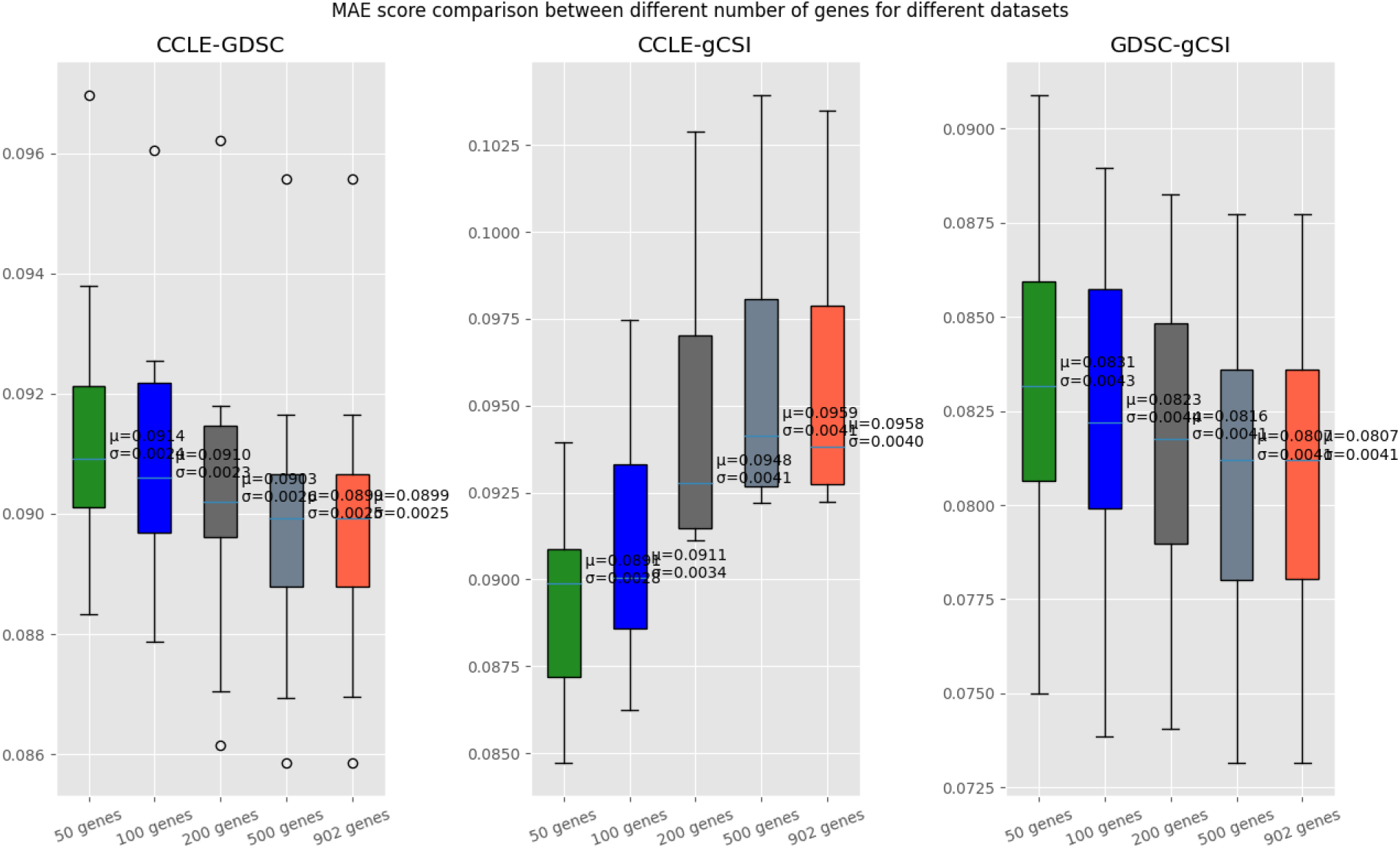
MAE after feature selection for different datasets

#### Task 3-Effect of the number of genes on model prediction

In this experiment, we wanted to see how robust our model was when different sources used different numbers of genes in their model for predicting drug response. We used mutual information to select 100 features from the 902 L1000 genes we used before. The results showed that for other approaches, feature selection (feature reduction) had an impact and improved their scores. However, for our method, the number of features did not matter significantly. Figure 6 shows the results after feature selection (compare with Figure 4). We used 10-fold cross-validation as before.

**Figure 6:**
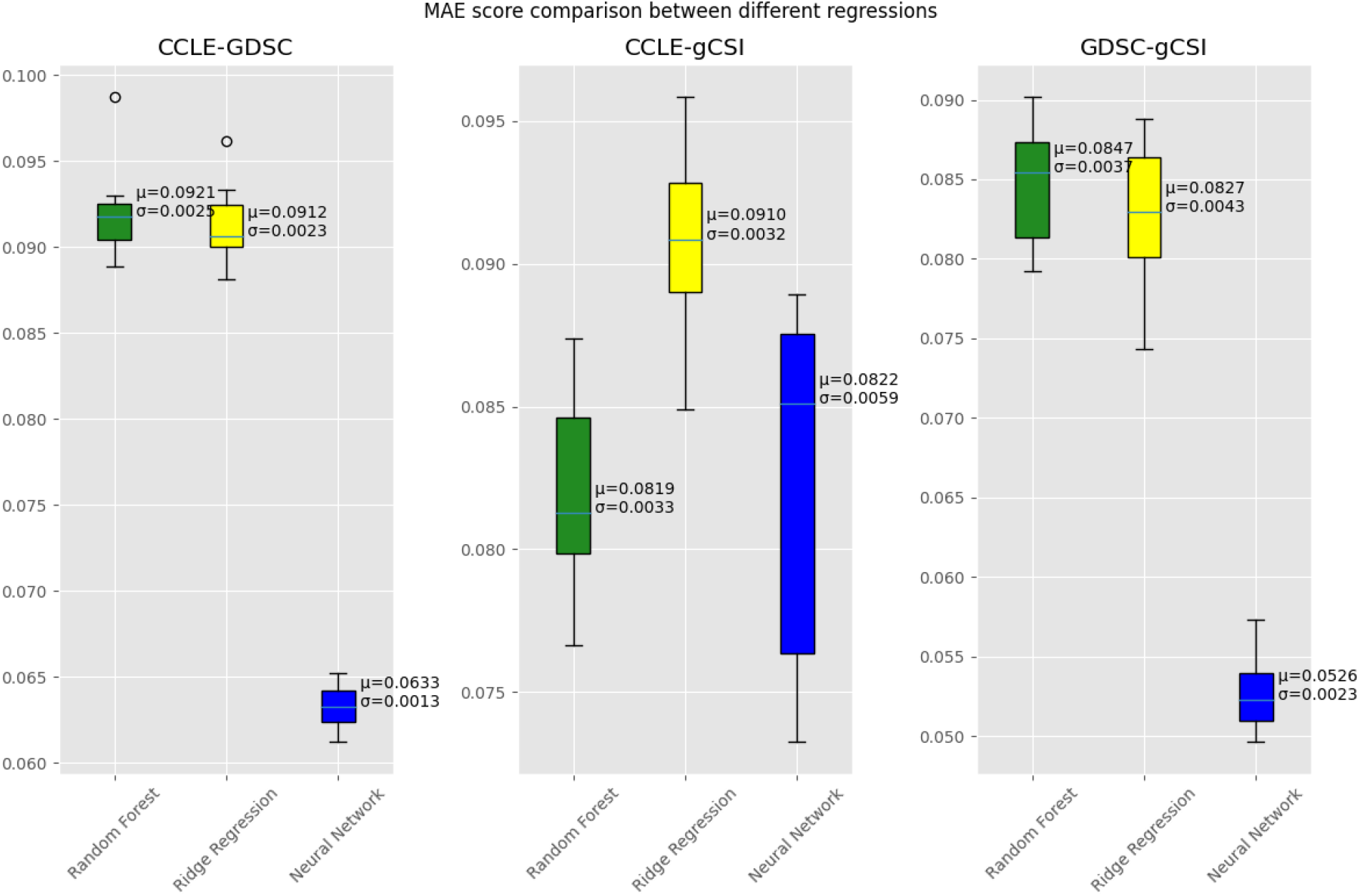
MAE for different regression techniques

#### Task 4-Effect of using different types of regression models for M1 and M2

We considered m1 and m2 as black boxes and replaced the ridge regression models with other regression models. Here we tried neural networks and random forest regression models. For random-forest regression, we set the max depth to 4 to prevent overfitting. Table 1 shows the results of the ablation study. For this part, we used a 70%-30% partition of the overlapping data for training and testing to save time.

## 3. Results

Since m1 and m2 as black box regression models, we used ridge regression models for both m1 and m2 in order to simplify the method in the comparison with baselines in Tasks 1-3. We further illustrated the performance of other ML/DL models in Task 4 to show the impact of the black box models.

### 3.1. Task 1-Performance on overlapping samples

Our analysis on three pairs of datasets (CCLE-GDSC, CCLE-gCSI, GDSC-gCSI) shows that our model outperforms SB, RA, CD and MA methods for all three pairs. Specifically, compared to other methods, our method achieved lower MAE scores of 0.090, 0.096, and 0.081 for the CCLE-GDSC, CCLE-gCSI, and GDSC-gCSI datasets respectively. However, for the CCLE-gCSI dataset, both the WA [MAE 0.0963] and MA [MAE 0.0938] methods perform as good as our method [MAE 0.0958] (Fig. 4). The combined dataset method performs the worst in both CCLE-GDSC [MAE 0.107] and CCLE-gCSI [MAE 0.107] datasets. The selected best method performed the worst in GDSC-gCSI [MAE 0.101] dataset.

### 3.2. Task 2-Effect of an increasing number of non-overlapping samples

There isn’t any substantial effect of increasing the number of non-overlapping samples on the result. Since our experiment is dependent on overlapping data, the change in non-overlapping samples does not make any significant difference in the result. For most datasets, the average MAE score is around 0.08 regardless of the size of non-overlapping data. We got a different result for the CCLE-GDSC dataset with changing CCLE compared to other datasets, with 2:1 ratio of CCLE:GDSC performing the best with the mean MAE score of 0.074. The overall experiment showed no other significant patterns in the results.

### 3.3. Task 3-Effect of the number of genes on model prediction

Our method performs better when the number of genes increases. This happens due to increased data for the model to learn from. For CCLE-GDSC [MAE 0.090] and GDSC-gCSI [MAE 0.081] datasets, our method performs the best when the maximum number of genes is used. On the contrary, CCLE-gCSI shows the opposite result as the maximum number of a gene gives the worst performance [MAE 0.96].

#### 3.3.1. Task 4-Effect of using different types of regression models for M1 and M2

Figure 6 shows the results of the ablation study. It is clear from the results that for all three pairs of datasets, the neural network model performs better than the other two models. For the CCLE-GDSC dataset, the neural network model produced an average MAE score of 0.063 whereas the random forest model had an average MAE of 0.092 and the ridge regression model had an average MAE of 0.091. Similarly, for the GDSC-gCSI dataset, the neural network model produced an average MAE score of 0.052 whereas the random forest model had an average MAE of 0.084 and the ridge regression model had an average MAE of 0.082. For the CCLE-gCSI dataset, the neural network and random forest method had an almost similar average MAE score of 0.082 whereas the ridge-regression method had an average MAE of 0.091.

## 4. Discussion

In this study, we developed a method for drug sensitivity prediction that draws on indirect information from several studies without utilizing shared data or models. The model outperformed other approaches that used multi-source data to predict medication response. This model was trained and evaluated in a preclinical setting that, to variable degrees, resembles patient treatment response. As a result, the model is applicable to both hospital settings and private pharmacogenomics datasets. To the best of the authors’ knowledge, we are the first to use a federated learning-like method for drug response prediction using multisource data. Our model performed better than methods in [21] and the traditional FL method when we only considered the overlapping samples from two datasets. Our model performed well since the knowledge from both sources is captured by the learned parameters during training. Also keeping in mind that paired test data might not always be available, our local model at one source did not require any external information from the other during testing. Unlike other FL methods, our method did not require model sharing and performed well by sharing only predicted training results in a dictionary format which did not disclose any private information. The result was robust when we used three different datasets pairwise for drug response prediction. We are novel in using drug structure, gene expression, and tissue type data for predicting drug response from multiple sources to build a generalized model. We also did extensive experiments to identify how generalizable the model is. Our simple but novel approach shows the utility of multiple data sources for building generalized models even when there are inconsistencies in data. While the proposed work is noteworthy, we also noticed that there are some limitations needed to be discussed in the proposed study:

Firstly, the proposed model leveraged the pairwise dataset instead of multiple studies to learn the inconsistency due to the constraint/assumption that there should be sufficient overlap among datasets for getting the best results. The number of overlapping samples decreased drastically with the increase in the number of datasets used, therefore, it resulted in a restriction on the number of datasets in training and testing (i.e., paired sets) in our study. We consider a more sophisticated mode that can learn the inconsistency from multiple datasets is needed in future work. Furthermore, our model was designed to handle the inconsistency based on the leverage of the overlapped information, which might not be satisfied in real scenarios where the overlapped information is not identified or mapped. For example, the inconsistency of patients’ demographic features (e.g., ages, gender), clinical features, and regimen plans might be diverse in different hospitals which are embedded in the EHR data [33]. Therefore, more studies are needed to improve the proposed model to leverage the multiple datasets with potential inconsistency but without a clear indication of the specific common overlapped information. Lastly, the proposed model targeted the drug response prediction based on the pharmacogenomic data in a pre-clinical setting where the locally trained models share an identical set of features. Since the proposed model does not require data sharing or model sharing among the local sets, the proposed model is robust to be adapted in other scenarios (e.g., EHR data in a clinical setting) where the mapped features are uncertain. However, since we did not study the cases of how the un-mapped features can impact the proposed model in the prediction tasks, we agreed that it may result in unsatisfactory results of the proposed model in such cases. A future study will be needed to investigate how the proposed model performs across multiple datasets with un-standard features or standard ones (e.g., ODSHI CDM [34] and PCORnet [35]).

## Data Availability

All data produced in the present study are available upon reasonable request to the authors

## 5. Acknowledgment

This work is supported by a grant from the National Institute of Health (NIH) NIGMS (R00GM135488).

